# The 8C model of vaccination readiness: A common framework to facilitate cross-study comparisons

**DOI:** 10.1101/2023.12.22.23300464

**Authors:** Piers Howe, Ferron Dearnley

## Abstract

Several factors can potentially influence an individual’s vaccination readiness. To facilitate cross-study comparisons, it is essential that researchers use a shared framework to measure these factors. This would not only help determine their relative importance cross different contexts but also would aid in tailoring interventions to enhance vaccine uptake. Historically, five psychological antecedents of vaccination were identified: confidence, complacency, constraints, calculation, and collective responsibility. This 5C scale was later expanded to a 7C model by incorporating two additional components: compliance and conspiracy. Building upon this framework, we propose an eighth component, certification, defined as the person’s self-report that, in the past, they have had to provide evidence of vaccination. This component addresses a significant gap in the 7C model, as some individuals reported taking the COVID-19 vaccine primarily to obtain proof of vaccination, a motivation not captured by the 7C model. Our confirmatory factor analysis (N = 406) of a bifactor model of US citizens’ self-reported COVID-19 vaccination status showed that this eighth component had good psychometric properties and the 8C model had slightly higher criterion validity than the 7C model. We present the 8C model as a framework that provides a richer and more complete descriptions of the factors that determine vaccination readiness and encourage future studies of vaccination readiness to utilise it.

## Introduction

Vaccines are the most effective intervention against a range of diseases, including COVID-19. While they do have risks, these risks are outweighed by the benefits. Despite this, there is growing hesitancy to be vaccinated, and this vaccine hesitancy has had substantial detrimental consequences. For instance, as of April 2023, there were at least 232,000 deaths in the USA due to COVID-19 among unvaccinated adults that could have been prevented by vaccinations (Jia et al., 2023). To put that number in perspective that is more than the number of US military fatalities in World War 1, the Korean War, the Vietnam War, the Gulf War and in the War on Terror combined (Statista, 2023).

To design effective interventions to increase vaccination uptake, the components of vaccination readiness must be better understood. We use the phrase “vaccination readiness” to refer to whether citizens are ready and willing to be vaccinated (Geiger et al., 2022). Prior work had argued that there are five psychological antecedents of vaccination and proposed a 5C scale of vaccine acceptance^1^ (Betsch et al., 2018). These antecedents were: *confidence* (the trust in the security and effectiveness of vaccinations), *complacency* (the tendency to avoid vaccinations due to the low perceived risk of infectious diseases), *constraints* (structural or psychological hurdles in daily life that make vaccination difficult or costly), *calculation* (the degree to which personal costs and benefits of vaccination are weighted), and *collective responsibility* (the willingness to get vaccinated to protect others). These factors have been shown to be predictive of vaccine uptake. For example, all five factors have been shown to be correlated with the uptake of the influenza vaccine and all factors other than *calculation* have been shown to be correlated with the uptake of the pneumococcal and shingles vaccines (Nicholls et al., 2021)

Subsequent work reconceptualised the scale as a bifactor model of vaccination readiness and argued for the addition of two further components: *compliance* (support for societal monitoring and sanctioning of people who are not vaccinated) and *conspiracy* (belief in conspiracies and false information related to vaccinations), resulting in a 7C model. The 7C model was shown to have greater criterion validity than the 5C model (Geiger et al., 2022).

Like the 5C model (Betsch et al., 2018), the 7C model is meant to provide a complete account of all the psychological antecedents of vaccination (Geiger et al., 2022). However, there is one other component of vaccination readiness that may be worth considering. Our initial scoping conversations suggested that some people agreed to be vaccinated against COVID-19 because they needed a vaccination certificate, either because their job required it or because they required it for some other purpose, such as to travel, visit a nursing home, go to a restaurant etc., and our preliminary exploratory study confirmed that that this was an important predictor of vaccination readiness. This reason is not addressed by the 7C model. As such it would seem sensible to consider adding to the 7C model an eighth component to represent this driver of vaccination.

### Current Research

Our objective was to make the existing 7C model more comprehensive by adding an additional component, *certification*, to create an 8C model. We defined *certification* as the person’s self-report that, in the past, they have had to provide evidence that they have been vaccinated. The main goal behind this addition wasn’t necessarily to enhance the model’s fit but rather to produce a more complete of vaccination readiness. This is essential as some individuals reported getting vaccinated mainly to have proof of their vaccination status, a reason that the 7C model overlooked. While previous research (Geiger et al., 2022) advocated for the 7C model over the 5C due to the 7C model having a superior fit, our study found that the criterion validity of the 8C model was only slightly better than that of the 7C model. However, our model offers a more holistic understanding of vaccination readiness in that it addresses a key psychological antecedent of vaccination that is ignored by the 7C model. To substantiate the validity of the added component, we conducted a confirmatory factor analysis to test the component’s psychometric properties and its effect on the overall model fit.

When Geiger et al. (2022) added the components *compliance* and *conspiracy* to the 5C scale to create the 7C bifactor model, they refined the existing components of the 5C scale to reduce overlap with the two new components and to improve the psychological characteristics of the combined scale. Because the 7C bifactor model is already well-designed and because the new component, *certification*, does not conceptually overlap with any of the existing components of the 7C model, we decided not to alter the 7C model beyond adding this eighth component to it.

Before settling on *certification*, we did consider other potential additions to the 7C model, which we derived both from theoretical considerations and conversations with experts and ordinary citizens. We identified eleven potential components and ran an exploratory study to test their psychometric properties. By considering their correlations with self-reported vaccination uptake, their correlations with the existing seven components of the 7C model, and their factor loadings in a bifactor model equivalent to the one utilised by Geiger et al. (2022), we eliminated ten of these potential components, leaving only *certification*. Details of the exploratory study, including the survey items, the raw data and the analysis (in R), can be found here: https://osf.io/8dcfr/. The exploratory study was pre-registered here: https://aspredicted.org/SNG_KLH

Because of the high communality of the seven components, Geiger et al. (2022) utilised a bifactor model, comprising a general factor of vaccination readiness and six specific nested factors for all components except for *confidence*, which served as a reference for the general factor. We adopted the same approach, except we added *certification* as a seventh specific nested factor. We ran a confirmatory factor analysis to determine how well this bifactor model fitted the data and compared the criterion validity of the 8C model to that of the 7C model. We found that the 8C model had greater criterion validity than the 7C model.

## Methods

### Sample and Procedure

Participants were recruited from Amazon Mechanical Turk (mturk.com). The study was approved by the University of Melbourne Office of Research Ethics and Integrity 2023-23370-38505-4. Participants were presented with an plain language statement and a consent form, and written consent was obtained before they were allowed to start the study. A pre-screener was run to identify bots and asked whether people self-reported as either fully vaccinated against COVID-19 or not. Excluding the bots, we then recruited 102 participants who reported not being fully vaccinated and 304 participants who reported being fully vaccinated against COVID-19, post exclusions. Participants were excluded if they did not finish the survey or if they “straight-lined” it. This study was pre-registered at https://aspredicted.org/9T2_V7J. The confirmatory study items, the pre-screener items, the confirmatory raw data and the analysis (in R) for the confirmatory study can be found here: https://osf.io/8dcfr/. Recruitment occurred 7-8 September, 2023. The mean age was 32.7 years (standard deviation 8 years). Seventy-six self-reported as female, 329 as male, 1 preferred not to say. With respect to education, two reported less than high school, 132 high school diploma, 13 reported some college but no degree, nine reported an associate degree (academic), four reported an associate degree (occupational), 178 reported a Bachelor’s degree, 62 reported a Master’s degree, five reported a Professional degree, one reported a Doctoral degree.

## Material

### Vaccination Readiness Scale

As a confirmatory factor analysis, we administered the full 8C scale with three items per component as shown in Table 1. Each item was rated on a 7-point Likert scale ranging from 1 = *strongly disagree* to 7 = *strongly agree*. In keeping with Geiger et al. (2022), we also studied a short version of the 8C scale, using just one item per component (bolded items in Table 1).

### Statistical Procedure

The factor loadings of the 8C bifactor model are shown in Figure 1. We evaluated model fit and factor saturation. As a rule of thumb, model fit is generally deemed acceptable at CFI and TLI ≥ .90, RMSEA < .08 and SRMR < .11, but there are circumstances where less good fits might be acceptable (Bentler, 1990; Hu & Bentler, 1999; Steiger, 1990). When predicting a dichotomous variable (e.g. willingness to vaccinate or self-reported vaccination status), the regressions were analysed using a logit link, and pseudo R^2^. All models were fitted using robust maximum likelihood estimation (MLR). The analysis was conducted in R (R Core Team, 2023) and Mplus Version 8 (Muthén & Muthén, 2017) using MplusAutomation Version 1.1.0 (Hallquist & Wiley, 2018) in R. The variances of the latent variables were fixed to one.

**Figure 1.**
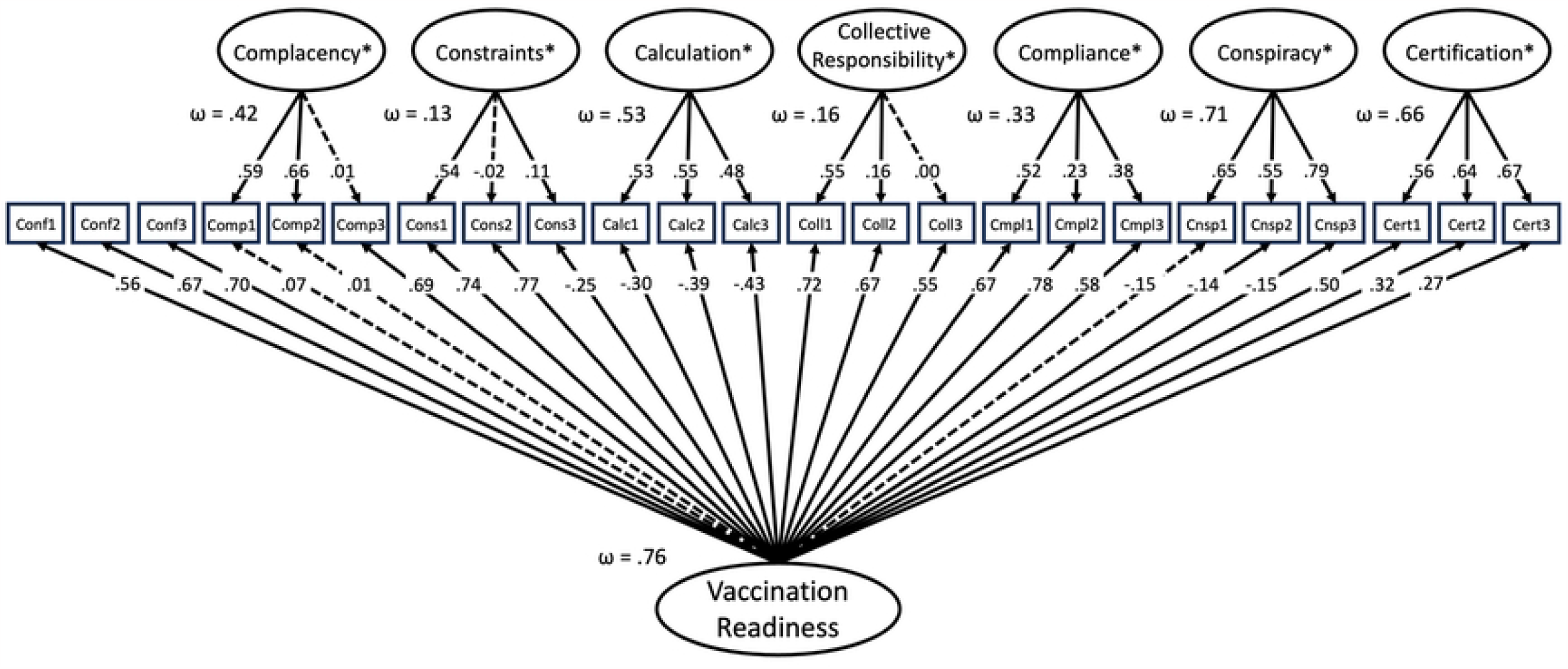

## Results

### Full Models

So that our findings could be readily compared to those of Geiger et al. (2022), we used a bifactor model with all items loading on the general factor of vaccination readiness. We did this both for the original 7C model and for the new 8C model. For both scales, all components except for confidence additionally loaded onto specific nested factors that represented variations in the components not explained by the general factor. For both models, confidence was chosen as the reference for the general factor, due to its high correlation with the general factor.

For the full 8C model, we found the following model fit values: CFI = 0.815, TLI = 0.786, RMSEA = .079, and SRMR = 0.096. The RMSEA and SRMR values are acceptable but the CFI and TLI values are a bit low (Bentler, 1990; Hu & Bentler, 1999; Steiger, 1990). We think this occurred because, to access sufficient numbers of unvaccinated participants, we used participants that we had not previously vetted, so the data quality may have been less than it otherwise would have been. Consistent with this suggestion, the fit parameters for the 7C model were similar to those for the 8C model: CFI = 0.821, TLI = 0.789, RMSEA = .083, and SRMR = 0.099.

As shown in Figure 1, the general factor loadings and the specific factor loadings for the three *certification* items were all positive. The factor saturation (McDonald’s ω; McDonald, 1999) for *certification* was of a similar magnitude to those of the other components (Table 1). The criterion validity for the 8C model was slightly more than that for the 7C model (0.687 vs 0.656).

Model fits can also be assessed by AIC and BIC. The 7C model had slightly lower values (AIC = 31449, BIC = 31746) than the 8C model (AIC = 35909, BIC = 36249). The 7C model was designed to provide a common framework to facilitate comparisons between different studies, by ensuring that everyone measured the same components (Geiger et al., 2022). As such, it includes components that may not be relevant to every study. The 8C model adopted the same philosophy. As such, we would not expect all components of the 8C model to be relevant in all situations. Including unnecessary components would likely increase the AIC and BIC values. To test for this, we constructed a 5C model that contained the five factors in the 8C model that loaded the highest on the general factor (vaccine hesitancy). These factors were *confidence, complacency, calculation, conspiracy* and *certification*. The AIC and BIC for this model were 22924 and 23132 respectively, showing that the 8C model can be reduced to a smaller model with lower AIC and BIC, if needed.

### Short Models

For the convenience of researcher, Geiger et al. (2022) also created a short version of the 7C scale, with one item per component. Following this lead, we created a short version of the 8C scale, selecting the *certification* item that loaded most onto the general factor.

For the short 8C, CFI = 0.905, TLI = 0.874, SRMR = 0.064, RMSEA = 0.085, AIC = 12287, BIC = 12379, and criterion validity of 0.314. For the short 7C, CFI = 0.898, TLI = 0.857, SRMR = 0.067, RMSEA = 0.096, AIC = 10797, BIC = 10877, and criterion validity of 0.296. For the short version of the previous 5C model, CFI = 0.867, TLI = 0.778, SRMR = 0.062, RMSEA = 0.102, AIC = 7849, BIC = 7905, and criterion validity of 0.306.

### Correlations and Reliability Analysis

A list of the correlations between the components can be found in Supplementary Table 1 here: https://osf.io/8dcfr/. *Certification* did not correlate highly with any other component (r ranged from -0.08 to 0.47). The individual certification items were moderately correlated with each other (r ranged from 0.45 to 0.52, Supplementary Table 2). The correlation between the full 8C scale and the short 8C scale was 0.892.

## Discussion

There are a large number of factors that can potentially affect vaccination readiness and vaccine uptake, and the importance of these components likely varies across different contexts (Betsch et al., 2018). It is therefore crucial that different studies use a common framework to measure these different components, so that the findings from different studies can be combined to determine how the relevance of the various components of vaccination readiness varies across contexts. Doing this will help researchers better tailor interventions for increasing vaccine uptake (WHO Regional Office for Europe, 2019).

The 7C model was designed to provide such a framework (Geiger et al., 2022). It attempted to include all the components that could affect vaccination readiness and vaccine uptake in different contexts. Although it is a powerful model that has been shown to have good psychometric properties (Geiger et al., 2022), it is missing a crucial component. In our preliminary investigations, we found that some people reported getting the COVID-19 vaccine to obtain proof of their vaccination status, which might need for their job, to travel, to visit relatives in a nursing, or even to eat in a restaurant. The 7C scale ignored this potential driver of vaccination uptake. We therefore added to the 7C scale an eighth component, *certification*, which we defined as the self-report of people that, in the past, they have needed to demonstrate evidence of vaccination.

During the COVID-19 pandemic, many governments required citizens to prove that they were vaccinated against COVID-19 before they were allowed to perform certain acts. While these restrictions were often justified as an attempt to reduce the spread of COVID-19, our conversations with experts and laypeople suggest that these restrictions were the primary reason why some people decided to get vaccinated. Since no existing component of the 7C model captures this driver of vaccination readiness, we argued that it was essential that *certification* be added to the 7C model, to create the 8C model.

To be clear, we didn’t expect adding an extra component to the 7C model would increase the model fit. Typically, models are constructed using the minimum number of components possible. However, the 7C model was constructed to include all the components that might, in different contexts, affect vaccination readiness. As such, it already contained a number of components that, while conceptually distinct, were correlated with each other. Thus, adding another component, especially if that component happened to be correlated with some of the existing components, would be unlikely to increase model fit.

Our confirmatory factor analysis showed that the psychometric properties of this additional component were very good, with all specific and general factor loadings both positive and of a reasonable magnitude. The general loadings ranged from 0.27 to 0.50, and the specific loadings ranged from 0.56 to 0.67. The McDonald’s ω was 0.66, which was comparable to the McDonald’s ω for the other six components which ranged from 0.13 to 0.71. Inter-item correlations ranged from 0.45-0.52. Its correlation with vaccination status (0.39) was the highest of any of the components. The model fit for the 8C model was very similar to that of the 7C model, except that the 8C model had slightly higher criterion validity.

Like Geiger et al. (2022), we also constructed a short version of the 8C scale by choosing the certification item with the highest general factor loading. The short 8C scale had better model fit than the full 8C scale. For the short 8C model, CFI = 0.91, TLI = 0.87, SRMR = 0.06 and RMSEA = 0.09. Conversely, for the full 8C model, CFI = 0.82, TLI = 0.79, SRMR = 0.10 and RMSEA = 0.08. The short 8C model had slightly better criterion validity than the short 7C model (0.31 vs 0.30). The correlation between the long and short 8C scales was 0.89.

A priori, one might be concerned that *certification* would overlap with *calculation*, which was defined as the degree to which personal costs and benefits of vaccination are weighted. If people were deciding to get vaccinated because they felt that they had to, you might expect them to score highly for both *certification* and *calculation*. If so, one would expect these two components to be highly correlated. This was not to be the case, with the correlation between *certification* and *calculation* being -0.08.

Although the labels appear superficially similar, *certification* and *compliance* are conceptually distinct. Whereas *certification* measures whether, in the past, a person needed to demonstrate evidence of vaccination, *compliance* measures the person’s support for societal monitoring and sanctioning of people who are not vaccinated. As such, the former refers to whether a person has previously been required to perform an action (i.e., prove that they have been vaccinated) whereas the latter measures the attitude of that person towards people who have not performed that action. So, while these two concepts are somewhat related, they are clearly conceptually different. This is reflected by them being somewhat correlated (r = 0.47).

There appears to be no reason to suspect that the *certification* would conceptual overlap with any of the remaining components of the 7C scale. For those components, correlations range from -0.08 to 0.39.

In conclusion, in this study we showed that it is necessary to add an eighth component, *certification*, to the 7C model. Because it does not conceptually overlap with any of the existing components, adding this component does not require us to alter the existing components of the 7C model in any way. Adding this extra component leaves the model fit essentially unchanged but does slightly increase the criterion validity. More importantly, there are strong theoretical reasons for adding this component: many people have reported that they got vaccinated because they needed vaccination certificates to partake in certain activities and mandating people have vaccination certificates before being allowed to partake in certain activities is one of the few ways governments can encourage vaccine uptake. Given a major driver behind this series of models is to help governments increase vaccine uptake (Betsch et al., 2018), not including this component would be inadvisable. That said, future work will need to determine how useful this extra component is in other settings. We note that in many countries children do need to prove that they have been vaccinated against certain diseases before being allowed to attend school and travellers are sometimes required to prove that they have been vaccinated against certain diseases before being allowed to enter a particular country. As such, we expect that *certification* will be predictive in other contexts as well.

## Data Availability

The survey items, the raw data and the analysis code (in R) can be found in a publicly accessible repository: https://osf.io/8dcfr/.

## Footnotes

1 While Betsch et al. (2018) refer to “vaccine acceptance", Geiger et al. (2022) refer to “vaccination readiness", to describe essentially the same construct. For the purpose of our study, we use the latter term as we employ the bi-factor model of Geiger et al. (2022).

